# Association of Physical Activity with the Incidence of Atrial Fibrillation in Persons >65 Years Old: The Atherosclerosis Risk In Communities (ARIC) Study

**DOI:** 10.1101/2021.12.07.21267411

**Authors:** Grace Fletcher, Aniqa B. Alam, Linzi Li, Faye L. Norby, Lin Y. Chen, Elsayed Z. Soliman, Alvaro Alonso

## Abstract

**Background:** Though moderate levels of physical activity (PA) seem to reduce the risk of atrial fibrillation (AF), the association of PA with AF in the elderly remains unclear.

**Methods:** We studied 5,166 participants of the Atherosclerosis Risk in Communities (ARIC) cohort that took part in visit 5 (2011-2013), were free of AF and had complete information on all variables. Self-reported PA was evaluated with a validated questionnaire and weekly minutes of leisure-time moderate to vigorous physical activity (MVPA) were calculated and categorized using the 2018 Physical Activity Guidelines for Americans (no activity [0 min/week], low [>0-<150 min/week], adequate [150-<300 min/week], high [≥300 min/week]). Incident AF between the visit 5 and the end of 2019 was ascertained from hospital discharges and death certificates. Cox models were used to calculate hazard ratios (HR) and 95% confidence intervals (CI) for AF by levels of physical activity adjusting for potential confounders.

**Results:** The mean (SD) age for the sample was 75 (5) years; 59% were female and 22% were Black. During a mean (SD) follow-up time of 6.3 (2.0) years, 703 AF events were identified. The association of MVPA with AF incidence showed a U-shaped relationship. Compared to those not engaging in MVPA, individuals with low MVPA had a 23% lower hazard of AF (HR= 0.77; 95% CI: 0.61, 0.96), while those with adequate MVPA had a 14% lower hazard (HR 0.86; 95% CI: 0.69, 1.06). High levels of MVPA were not associated with AF risk (HR 0.97; 95% CI: 0.78, 1.20). There was no evidence of heterogeneity when stratified by race and sex.

**Conclusion:** This study suggests that being involved in low to moderate levels of MVPA was associated with a reduced hazard of AF. There was no evidence of increased risk of AF in those with higher levels of MVPA.

## INTRODUCTION

Atrial fibrillation (AF) is one of the most common cardiac arrhythmias found in developed countries.^1^ Older age, as well as the presence of cardiovascular risk factors such as hypertension, obesity and diabetes, have been consistently linked to an increased risk of being diagnosed with AF.^2^ Lifestyles may also play a role in the development and prevention of AF.^3^ In particular, multiple studies have evaluated the impact that physical activity (PA) has in the development of AF. Meta-analyses of these studies suggest a U-shaped association between PA and AF risk, with lower risk among individuals engaged in moderate levels of physical activity but no benefit with higher levels, compared to those with low levels.^4, 5^ In fact, individuals engaging in high-endurance PA (e.g. long-distance runners and skiers, professional cyclists) seem to have a higher risk of developing AF.^6^ These observations suggest a complex relationship of PA levels and intensity with AF risk.

AF risk is particularly high among persons 65 and older. The evidence on the effects of PA on AF risk in this age group, for which preventive interventions would have the highest impact, is limited. An analysis of the Cardiovascular Health Study, including 5446 persons older than 65 (mean age 73), found an inverse linear association between leisure time PA and AF risk, but a U-shaped association between exercise intensity and AF, with lower AF risk associated with moderate-intensity exercise but not with high-intensity exercise.^7^ Additional studies on the association of PA with AF risk in older individuals, particularly among racially diverse populations, are needed to inform preventive guidelines in this high-risk population.

Thus, to address existing limitations of the literature and provide additional evidence that could inform preventive recommendations in older individuals, we evaluated the association of leisure-time PA with the incidence of AF in White and Black persons older than 65 years of age enrolled in the Atherosclerosis Risk in Communities (ARIC) study, a prospective cohort in the United States.

## METHODS

### Study Population

The ARIC study is a prospective cohort that recruited people 45-64 years of age from 4 communities in the Unites States (Forsyth County, North Carolina; Jackson, Mississippi; Washington County, Maryland; and selected suburbs of Minneapolis, Minnesota) in 1987-1989.^8^ Participants had subsequent study visits in 1990-1992, 1993-1995, 1996-1998, 2011-2013, 2016-2017, and 2018-2019. For this particular analysis, we included participants attending the 2011-2013 (visit 5). We excluded participants with prevalent AF at that visit, those reporting race other than White or Black, as well as non-White participants from the Minnesota and Washington County sites (due to small numbers), and those with missing information on PA or other study covariates (**Figure 1**). The Institutional Review Boards at each participating ARIC study center approved this study, and all participants provided written informed consent.

**Figure 1.**
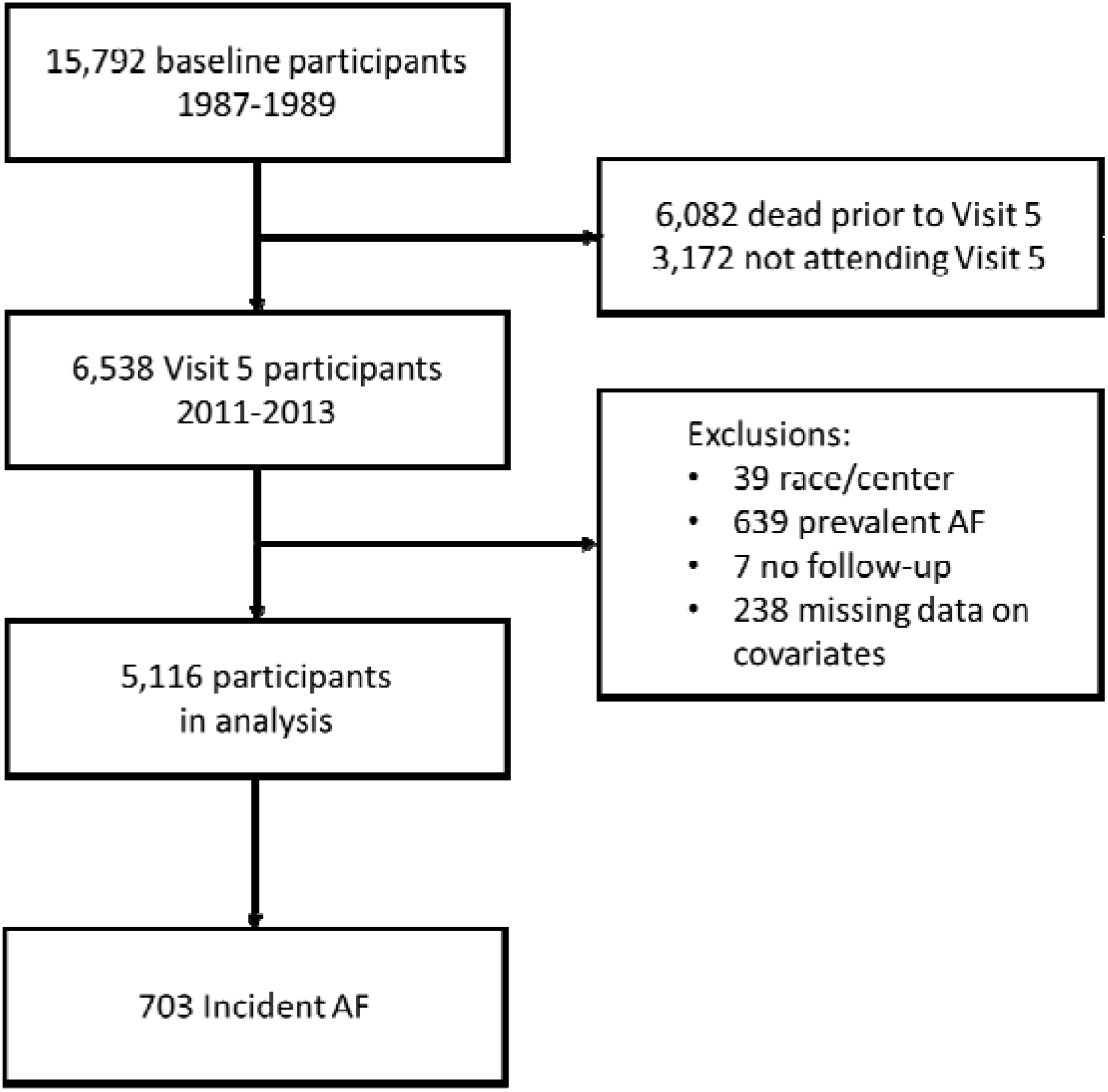
Study inclusion flowchart, Atherosclerosis Risk in Communities study. AF: atrial fibrillation

### Assessment of Physical Activity

Physical activity was measured with a modified Baecke Physical Activity Questionnaire. The questionnaire asks participants about their leisure, work and sports-related PA over the last year.^9^ Specifically, the questionnaire asks for up to four of the most commons sports or leisure-time activities, as well as the duration and frequency. This information was used to calculate the time (in minutes/week) and intensity (in metabolic equivalent of task [MET] minutes per week) of each activity, and added to obtain weekly levels of moderate to vigorous physical activity (MVPA).^10^

The primary exposure in this study was defined using categories of time spent on MVPA per week according to recommendations outlined by the 2018 Physical Activity Guidelines for Americans.^11^ Specifically, participants were classified in the following groups: no MVPA (reference), <150 min/week of moderate physical activity and <75 min/week of vigorous physical activity (low MVPA), 150-299 min/week of moderate physical activity or 75-149 min/week of vigorous physical activity or an equivalent combination of MVPA (recommended MVPA), and ≥300 min/week of moderate physical activity or ≥150 min/week of vigorous physical activity or an equivalent combination of MVPA (high MVPA). To combine vigorous and moderate PA, 1 minute of vigorous activity was counted as 2 minutes of moderate activity. Thus, someone engaging in 100 min/week of moderate PA and 30 min/week of vigorous PA, would be included in the recommended MVPA category (100 + 30 × 2 = 160 min/week MVPA).

### Ascertainment of Incident Atrial Fibrillation

The main outcome was incident AF between visit 5 and end of 2019 among those without previously diagnosed AF (through the end of 2017 for participants in the Jackson site). AF was identified from electrocardiograms done at study visits 1 through 5, hospital discharge codes ICD-9-CM 427.3x and ICD-10-CM I48.x not occurring in the context of cardiac surgery, and from death certificates with AF as underlying or contributing cause of death (ICD-10 I48).^12^

### Other Covariates

Age, sex, race, education, smoking (never, former, current) and alcohol (never, former, current) use were self-reported. Height and weight were measured at the time of the study visit and used to calculate body mass index (BMI). Blood pressure was measured three times and the second and third measurements were averaged to calculate systolic and diastolic blood pressure. Hypertension was defined as systolic blood pressure ≥140 mmHg or diastolic blood pressure ≥90 mmHg or use of antihypertensive medication. Blood glucose was measured in samples obtained during the visit. Diabetes was defined as a fasting blood glucose ≥126 mg/dL, non-fasting blood glucose ≥200, use of antidiabetic medication or a self-reported physician diagnosis of diabetes. Prevalent coronary heart disease and heart failure were based on prior history of these conditions using ARIC-specific definitions, as described elsewhere.^13, 14^ Physical function was evaluated using the Short Physical Performance Battery (SPPB), an assessment tool for evaluation of lower extremity functioning in older adults, ranging from 0 (worst) to 12 (best function). ^15^ We calculated the underlying risk of AF using the Cohorts for Heart and Aging Research in Genomic Epidemiology (CHARGE)-AF score, which provides the 5-year risk of AF based on age, race, height, weight, systolic and diastolic blood pressure, smoking, antihypertensive medication use, diabetes, and history of coronary heart disease and heart failure.^2^

### Statistical Analysis

Statistical analyses were conducted using SAS 9.4 statistical software. In the primary analysis, physical activity was categorized using MVPA cutoffs described above. Additionally, we explored associations of AF with vigorous and moderate physical activity. Categories for vigorous physical activity were 0, >0-74, and ≥75 min/week and for moderate physical activity were 0, >0-149, ≥150-299, and ≥300 min/week. Cox regression models were used to calculate hazard ratios (HR) and 95% confidence intervals (CI) of AF by levels of MVPA. An initial model adjusted for age, sex, race/center, and education. A second model additionally adjusted for alcohol drinking, cigarette smoking, diabetes, hypertension, prevalent coronary heart disease, prevalent heart failure, BMI, and SPPB. We conducted additional analysis stratifying by race, sex, and the underlying risk of AF, as calculated by the CHARGE-AF score.^2^ We tested the proportional hazards assumption including interaction terms between exposure and time. No evidence of lack of proportionality in the hazards was found.

## RESULTS

The mean (SD) age for the 5166 eligible participants was 75 (5) years; of them, 59% were female and 22% were Black. During a mean (SD) follow-up time of 6.3 (2.0) years, 703 AF events were identified. **Table 1** presents patient characteristics by MVPA category (33% no MVPA, 19% low MVPA, 25% recommended MVPA, 23% high MVPA). Participants engaging in higher levels of MVPA were less likely to be female, Black, and to have a prior history of hypertension, diabetes, or heart failure. Age and predicted risk of AF according to the CHARGE-AF score were similar across MVPA categories.

**Table 1.**
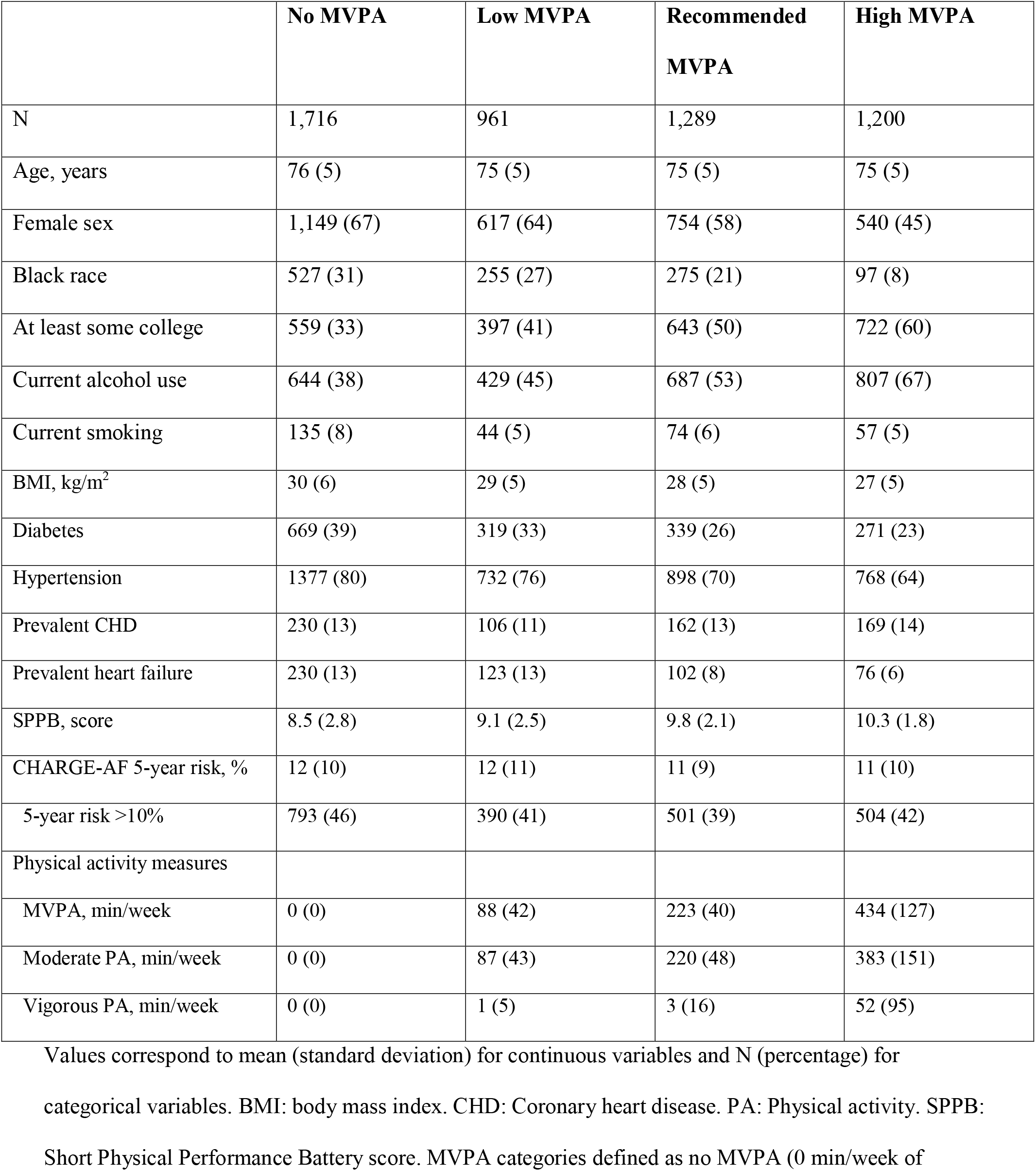
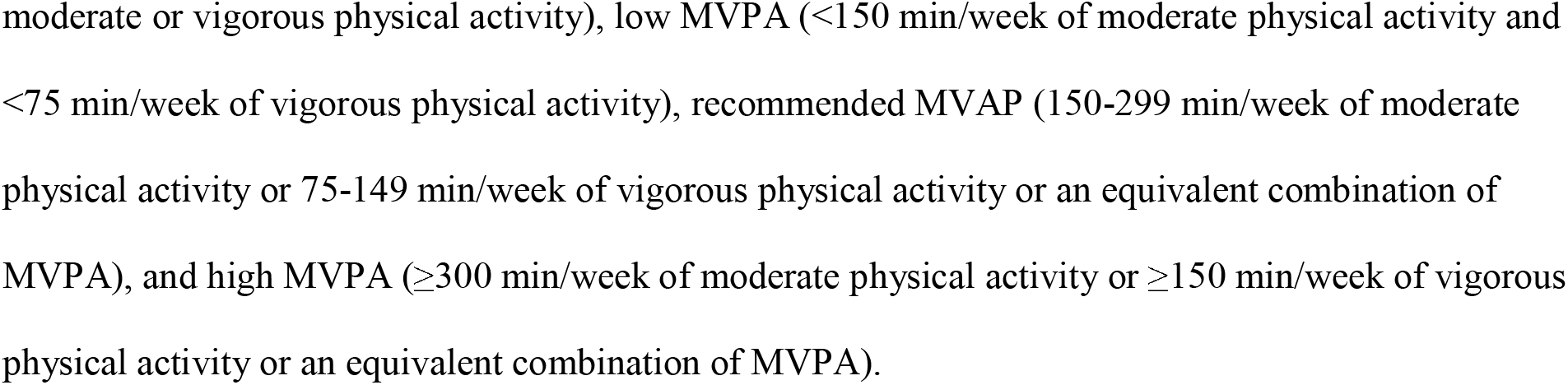
Baseline characteristics of study participants by levels of moderate to vigorous physical activity (MVPA). Atherosclerosis Risk in Communities study, 2011-13.

|  | **No MVPA** | **Low MVPA** | **Recommended MVPA** | **High MVPA** |
| --- | --- | --- | --- | --- |
| N | 1,716 | 961 | 1,289 | 1,200 |
| Age, years | 76 (5) | 75 (5) | 75 (5) | 75 (5) |
| Female sex | 1,149 (67) | 617 (64) | 754 (58) | 540 (45) |
| Black race | 527 (31) | 255 (27) | 275 (21) | 97 (8) |
| At least some college | 559 (33) | 397 (41) | 643 (50) | 722 (60) |
| Current alcohol use | 644 (38) | 429 (45) | 687 (53) | 807 (67) |
| Current smoking | 135 (8) | 44 (5) | 74 (6) | 57 (5) |
| BMI, kg/m<sup>2</sup> | 30 (6) | 29 (5) | 28 (5) | 27 (5) |
| Diabetes | 669 (39) | 319 (33) | 339 (26) | 271 (23) |
| Hypertension | 1377 (80) | 732 (76) | 898 (70) | 768 (64) |
| Prevalent CHD | 230 (13) | 106 (11) | 162 (13) | 169 (14) |
| Prevalent heart failure | 230 (13) | 123 (13) | 102 (8) | 76 (6) |
| SPPB, score | 8.5 (2.8) | 9.1 (2.5) | 9.8 (2.1) | 10.3 (1.8) |
| CHARGE-AF 5-year risk, % | 12 (10) | 12 (11) | 11 (9) | 11 (10) |
| 5-year risk >10% | 793 (46) | 390 (41) | 501 (39) | 504 (42) |
| Physical activity measures |  |  |  |  |
| MVPA, min/week | 0 (0) | 88 (42) | 223 (40) | 434 (127) |
| Moderate PA, min/week | 0 (0) | 87 (43) | 220 (48) | 383 (151) |
| Vigorous PA, min/week | 0 (0) | 1 (5) | 3 (16) | 52 (95) |

Values correspond to mean (standard deviation) for continuous variables and N (percentage) for categorical variables. BMI: body mass index. CHD: Coronary heart disease. PA: Physical activity. SPPB: Short Physical Performance Battery score. MVPA categories defined as no MVPA (0 min/week of
moderate or vigorous physical activity), low MVPA (<150 min/week of moderate physical activity and <75 min/week of vigorous physical activity), recommended MVAP (150-299 min/week of moderate physical activity or 75-149 min/week of vigorous physical activity or an equivalent combination of MVPA), and high MVPA ( $\geq 300$ min/week of moderate physical activity or $\geq 150$ min/week of vigorous physical activity or an equivalent combination of MVPA).

**Table 2** shows the association between MVPA levels and the incidence of AF. There was an overall U-shaped association, with the highest rates in those not engaging in MVPA, lowest among those with low or recommended levels of MVPA, and higher again in those with high MVPA. In the analysis adjusted for demographics, lifestyles and clinical variables, participants engaging in low levels of MVPA had a 23% lower hazard of AF compared to those in the no MVPA category (HR 0.77; 95% CI: 0.61, 0.96). The corresponding figure for those in the recommended MVPA category was 14% (HR 0.86; 95% CI: 0.69, 1.06). Participants engaging in high levels of MVPA had a risk of AF similar to those with no MVPA (HR 0.97; 95% CI: 0.78, 1.21). Associations were similar in subgroups defined by sex, race, and baseline AF risk, as defined by the CHARGE-AF score (**Table 3**).

**Table 2.** Association of moderate to vigorous physical activity (MVPA) levels with incidence of atrial fibrillation (AF). Atherosclerosis Risk in Communities study, 2011-2019.

|  | **No MVPA** | **Low MVPA** | **Recommended MVPA** | **High MVPA** |
| --- | --- | --- | --- | --- |
| AF events | 268 | 112 | 156 | 167 |
| Person-years | 10,338 | 5,980 | 8,323 | 7,933 |
| IR (95% CI)* | 26 (23, 29) | 19 (15, 23) | 19 (16, 22) | 21 (18, 24) |
|  | HR (95% CI) |  |  |  |
| Model 1 | 1 (ref.) | 0.73 (0.59, 0.91) | 0.73 (0.59, 0.89) | 0.80 (0.65, 0.99) |
| Model 2 | 1 (ref.) | 0.77 (0.61, 0.96) | 0.86 (0.69, 1.06) | 0.97 (0.78, 1.21) |

\* Incidence rate per 1,000 person-years.
AF: atrial fibrillation. CI: Confidence interval. HR: Hazard ratio. IR: Incidence rate.
Model 1: Cox proportional hazards model adjusted for age, sex, race/center and education.
Model 2: Cox proportional hazards model adjusted for Model 1 variables and alcohol drinking, cigarette smoking, diabetes, hypertension, prevalent coronary heart disease, prevalent heart failure, body mass index, and Short Physical Performance Battery score.

**Table 3.** Hazard ratios and 95% confidence intervals of atrial fibrillation (AF) for categories of moderate to vigorous physical activity (MVPA) by sex, race, and baseline risk of AF. Atherosclerosis Risk in Communities study, 2011-2019.

|  | **No MVPA** | **Low MVPA** | **Recommended**<br>**MVPA** | **High MVPA** | P for interaction |
| --- | --- | --- | --- | --- | --- |
|  | HR (95% CI) |  |  |  |  |
| **Sex** |  |  |  |  |  |
| Female | 1 (ref.) | 0.76 (0.57, 1.01) | 0.89 (0.67, 1.17) | 0.89 (0.64, 1.23) | 0.82 |
| Male | 1 (ref.) | 0.76 (0.53, 1.08) | 0.83 (0.60, 1.14) | 1.03 (0.76, 1.40) |  |
| **Race** |  |  |  |  |  |
| Black | 1 (ref.) | 0.57 (0.32, 1.04) | 0.70 (0.39, 1.27) | 0.48 (0.17, 1.38) | 0.49 |
| White | 1 (ref.) | 0.80 (0.63, 1.02) | 0.89 (0.71, 1.11) | 1.02 (0.81, 1.28) |  |
| **Baseline AF**<br>**risk** |  |  |  |  |  |
| <10% | 1 (ref.) | 0.80 (0.56, 1.15) | 0.86 (0.62, 1.20) | 0.78 (0.54, 1.15) | 0.20 |
| ≥10% | 1 (ref.) | 0.74 (0.56, 0.99) | 0.83 (0.63, 1.09) | 1.06 (0.82, 1.38) |  |

Cox regression models adjusted for age, sex and race/center (if applicable), education, alcohol drinking, cigarette smoking, diabetes, hypertension, prevalent coronary heart disease, prevalent heart failure, body mass index, and Short Physical Performance Battery score.

We also evaluated separately the association of moderate and vigorous PA with the incidence of AF (**Figure 2**). Moderate PA showed an L-shaped association with AF risk, with the highest risk among those not participating in any moderate PA, and the lowest in those participating in >0-<150 min/week of moderate PA. There was no evidence of increased risk of AF in participants engaged in ≥300 min/week. The association of vigorous PA with AF risk was U-shaped, with the lowest risk in those participating in >0-<75 min/week, and similar risk in those with 0 and ≥75 min/week.

**Figure 2.**
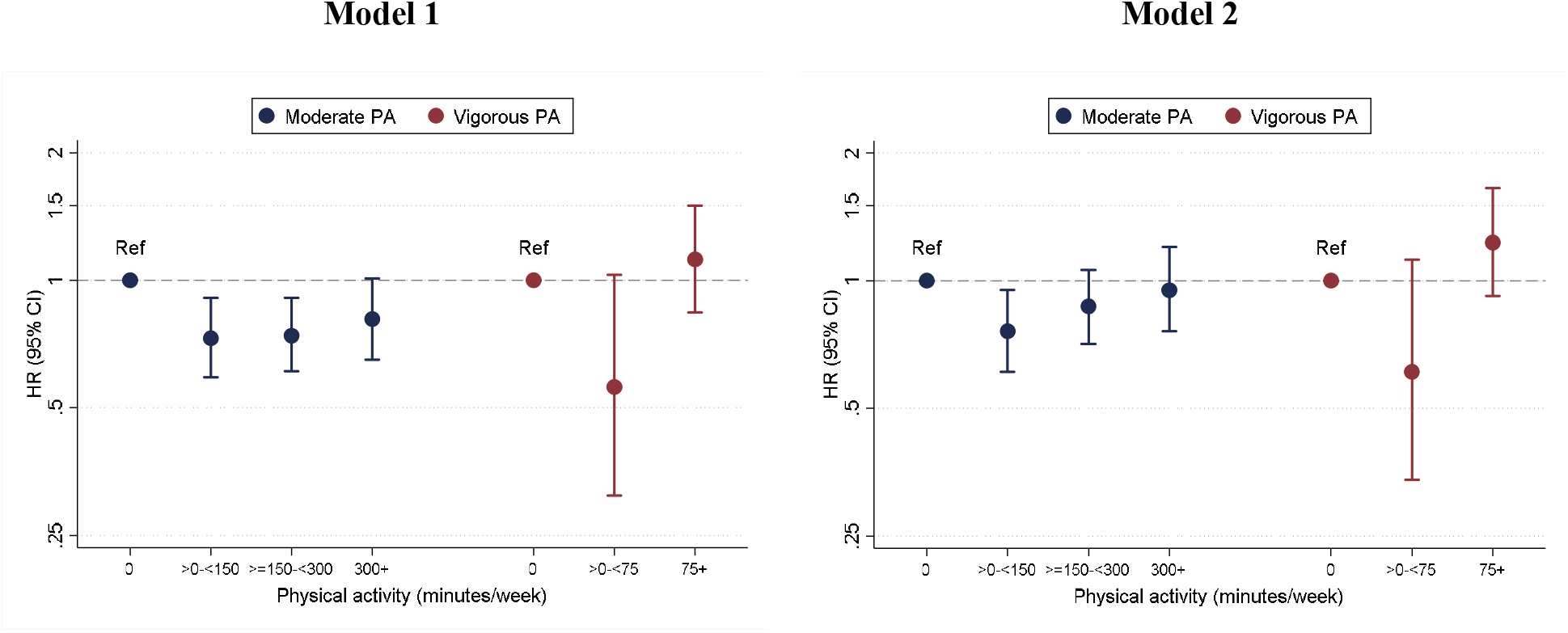
Hazard ratios and 95% confidence intervals of atrial fibrillation (AF) for categories of moderate and vigorous physical activity (PA). Model 1 is a Cox regression model including moderate and vigorous PA categories in the same model adjusted for age, sex, race/center, and education. Model 2 additionally adjusts for education, alcohol drinking, cigarette smoking, diabetes, hypertension, prevalent coronary heart disease, prevalent heart failure, body mass index, and Short Physical Performance Battery score. Atherosclerosis Risk in Communities study, 2011-2019.

## DISCUSSION

In this large prospective community-based study of individuals older than 65, we observed that engaging in some degree of MVPA was associated with a reduced risk of developing AF, with less or no benefit among those reporting very high levels of MVPA. The associations were similar across groups defined by sex, race, or underlying AF risk. The beneficial association was observed for intermediate levels of moderate PA and vigorous PA separately, though the limited number of study participants engaging in vigorous PA resulted in imprecise estimates of association. Overall, our findings suggest that engaging even in small amounts of moderate or vigorous PA could reduce the risk of AF among persons 65 and older, and that the risk of AF is not particularly elevated among those with higher levels of PA.

These results from the ARIC cohort are mostly consistent with those reported in other studies. A recent dose-response meta-analysis of the literature including data from 15 studies and over 1.4 million individuals (median age 55 years) also described a U-shaped association, with the lowest risk among individuals engaging in 1000-1500 MET-minutes per week.^5^ Few studies have specifically focused on persons 65 and older. The Cardiovascular Health Study, which followed 5,446 participants 65 and older for 12 years, found that amount of leisure-time physical activity had an inverse linear association with risk of AF (HR, 0.64; 95% CI, 0.52-0.79, comparing 5^th^ to 1^st^ quintile), while the association between exercise intensity and AF risk was U-shaped, with HR (95% CI) of AF compared to no exercise of 0.85 (0.69-1.03) for low intensity, 0.72 (0.58-0.89) for moderate intensity, and 0.87 (0.64-1.19) for high intensity exercise.^7^

AF risk seems to be higher in athletes and other individuals engaged in intense endurance exercise, particularly younger ones.^16^ In a meta-analysis of 13 studies, including 6,816 athletes and 63,662 controls, the odds of AF were 2.5 higher in athletes compared to controls (OR 2.5; 95% CI: 1.7-3.5), with the association between being an athlete and AF risk weakening with advancing age.^16^ However, average levels of PA among participants in community-based studies, including ARIC, are generally much lower than those including athletes, making both types of studies difficult to compare.

Engaging in PA could contribute to a reduced risk of AF through multiple mechanisms. PA plays a key role in the prevention and management of several risk factors for AF, including elevated blood pressure, obesity, diabetes, and other cardiovascular diseases (heart failure, coronary heart disease.)^17^ By preventing these risk factors, higher levels of PA would result in decreased AF risk over time. We observed, however, that the beneficial association was even present among ARIC participants with a high underlying risk of AF, due in part to presence of AF risk factors. PA could also impact AF risk by directly affecting the atrial substrate responsible for AF development. On the one hand, higher levels of PA in older individuals have been associated with better systolic and diastolic function, in turn preventing the development of the AF atrial substrate.^18^ On the other hand, very high levels of PA could lead to atrial enlargement (but not to atrial mechanical function),^19^ atrial ectopy, atrial fibrosis, increased vagal tone, and changes in electrolytes that can trigger AF or contribute to the development of a susceptible cardiac substrate.^20, 21^ The balance between these two opposing pathways may explain the observed U-shaped association between PA and AF risk.

Our findings have relevance for public health guidelines and individual preventive medicine recommendations, by reinforcing current PA guidelines for persons 65 and older and demonstrating absence of harm at those recommended levels in relation to AF risk, even among individuals at higher risk of developing AF. Specifically, the current recommendation to do 150-300 minutes a week of moderate-intensity aerobic PA or 75-150 minutes of vigorous-intensity aerobic PA or an equivalent combination was attainable by a large proportion of our study population (close to 50% were in this category or higher) and was associated with a small reduction in AF risk. Concerns about risk of developing AF should not preclude engaging in moderate levels of PA given the multiple benefits of exercise in the prevention of cardiometabolic diseases and other aging-associated disorders.

The validity of our results is supported by the robust follow-up of the ARIC cohort, with minimal losses to follow-up, the richness of information on relevant covariates, allowing for confounding control, and the diversity of the study population, which facilitated the evaluation of the association separately in males and females, and in Black and White Americans. Some limitations need to be acknowledged, however. Information on PA was self-reported and we did not have objectively measured PA, which is likely to lead to measurement error, non-differential with regards to the study endpoint. AF ascertainment is potentially incomplete, missing subclinical AF as well as AF exclusively managed in an outpatient setting in participants not being hospitalized during their follow-up. This lack of sensitivity in case ascertainment is also likely to be non-differential with regards to PA. Validation studies for the PA assessment and the method of AF ascertainment in ARIC minimize these concerns.^12, 22^ Finally, by design, Black participants in the ARIC cohort were recruited primarily at the Jackson site, limiting our ability to separate the effects of race and geographic location.

In conclusion, we observed a U-shaped association between MVPA and AF risk in older persons. These findings support current PA recommendations for this age group and can be used to reassure individuals concerned about AF risk associated with higher levels of MVPA. Future work should aim to understand mechanisms through which PA influences AF risk in this age group, to identify individuals that may particularly benefit from increased levels of PA, and to develop interventions that could reduce the risk of AF in high-risk individuals.

## Data Availability

ARIC data can be obtained through the NHLBI BioLINCC repository or by request to the ARIC Coordinating Center at UNC-Chapel Hill.

## ACKNOWLEDGEMENTS

The authors thank the staff and participants of the ARIC study for their important contributions.

